# Kinetics of Immunoglobulins in Septic Shock Patients Treated with an IgM and IgA-Enriched Intravenous Preparation: an Observational Study

**DOI:** 10.1101/2020.09.08.20189183

**Authors:** Giorgio Berlot, Alice Scamperle, Tatiana Istrati, Roberto Dattola, Irene Longo, Antonino Chillemi, Silvia Baronio, Giada Quarantotto, Erik Roman-Pognuz, Ariella Tomasini, Mattia Bixio

## Abstract

**Objective:** to assess the variations of the blood levels of immunoglobulins (Ig) in septic shock patients treated with an Ig preparation enriched in IgM and IgA (eIg).

**Design:** The blood levels of Ig in survivors (S) and non-survivors (NS) of a group of septic shock patients were measured before the initial administration (D0) and one (D1), four (D4) and seven (D7) days thereafter. The SAPS II score, the capillary permeability, the primary site of infection, the antibiotic appropriateness, the microorganisms isolated and the outcome at 28 days were also assessed.

**Results:** In the interval D0-D7 the IgM increased significantly only in the S while remained stable in NS; the IgA significantly increased in both groups; the IgG did not vary significantly in both groups. At D7 the capillary permeability significantly decreased in S but not in NS.

**Conclusions:** The kinetics of the different classes of Ig after eIg were different between S and NS. This could be related either to (a) different capillary permeability in the two groups and/or (b) higher Ig consumption in NS. Further studies to confirm the benefits of eIg in the treatment of sepsis syndrome, to define the specific target population and the correct eIg dose are warranted.

## Introduction

Despite different studies and meta-analyses demonstrated a better outcome in septic patients who received intravenous immunoglobulins (IvIg)^1–3^ compared with controls, the Surviving Sepsis Campaign’s guidelines (SSC) recommend against their use due to the lack of trials fulfilling the Evidence-Base Medicine criteria^4^. The main criticisms are based on (a) patients-related variables, including the number and severity of comorbidities, the underlying immunitary capabilities and the adequacy of other concomitant therapies; (b) the presence and the amount of antibodies against the responsible germ(s), including Multiple Drug Resistant (MDR) strains in the preparation administered; and, finally, (c) the class of IvIg used^4, 5^. Currently, just one preparation is enriched with elevated concentrations of IgM and IgA (eIg) (12% each) whereas the other ones contain these Ig in trace amounts only.

As far as this latter issue is concerned, it appears that the positive effects of IvIg are more marked in patients treated with eIg than with IgG only^6–8^. Different factors could account for this finding, including the IgA-associated protective effects on the gut mucosa^9–11^ and the pentameric structure of the IgM that allows their binding to different antigens located on the germ surface or to pathogens-associated molecular patterns (PAMP) with the subsequent activation of the complement^12,13^; moreover, IgM molecules exert an immumodulatory effect by scavenging excessive complement factors and blunting the production of some sepsis mediators^14,15^.

Besides the above listed confounding factors, another critical point associated with the IvIg is represented by the target blood values to achieve^16,17^. This could be particularly relevant in the case of eIg due to the high cost of the only preparation available. Actually, the manufacturer’s indications recommend a standard dose of 250 mg/kg of body weight for three consecutive days and the choice to add further doses or not is left to the physician, independently from the initial and end-treatment blood values of the γ-globulins. This approach carries the inherent risk of over- or under-administration of the eIg and is far from the concept of precision medicine.

As some investigations demonstrated that in different critical conditions the kinetics of endogenous Ig was associated with the outcome^16,17^, we measured the time course of these substances in survivors and non-survivors of a group of septic shock patients before, during and after the administration of eIg.

### Patients and Methods

This is a multicenter, observational, non-interventional study, involving two general adult Intensive Care Units (ICU) (the Department of Anesthesia and Intensive Care of Trieste and the Department of Anesthesia and Intensive Care of Alessandria). eIg (Pentaglobin®, Biotest, Dreiech, Germany) are routinely used in these ICUs and the study did not imply any interventions other than the standard medical care, the local Ethical committees considered the patients’ informed consent unneeded.

We included 54 patients admitted with septic shock in both ICUs. Septic shock was defined according to SEPSIS 3 criteria^18^.

The overall treatment followed the SSC guidelines. Exclusion criteria were age < 18 years, previous administration of IvIg, hematological tumors, known immune depression (i.e. AIDS), recent or ongoing treatment with immunosuppressant and a life expectancy < 3 months. The eIg were given at the dose of 250 mg/kg/day; the infusion lasted 10 h and was repeated for 3 days (total dose 750 mg/kg. For all patients, we collected demographic characteristics, diagnosis at admission and type of admission (surgical or medical). We used the SAPS score to assess the severity of the condition of each patient. We obtained blood samples for γ-globulins measurement immediately before starting the eIg infusion (D0) and successively after one (D1), four (D4), and seven (D7) days. The capillary leak index (CLI), calculated according to the formula: C-reactive protein (CRP) (milligrams/100 ml) / albumin (grams/liter) * 100 was used as a marker of capillary permeability at D0 and D4.

The primary site of infection, the antibiotics administered and their appropriateness, the microorganisms isolated, and the outcome at 28 days were also recorded.

The statistical analyses were performed using GraphPad Prism Version 8.0.2 (GraphPad Software, La Jolla, CA, http://www.graphpad.com) and R statistical package, software version 3.3.3. Descriptive statistics were analyzed for all the variables. Discrete variables were expressed as percentage and continuous variables as mean ± SD or median (25th-75th percentiles). Categorical variables were compared by Chi-square or Fisher exact test. Continuous variables were compared using Student *t* test. The variation of the blood levels of Ig was expressed as percentage and nonparametric tests, such as Mann-Whitney, were used since the data were not normally distributed. A level of *P value* less than 0.05 was considered statistically significant.

## Results

Overall, 54 patients were enrolled (Table 1); non-survivors at 28 days (NS = 14) had a significantly higher SAPS 2 score as compared with survivors (S = 40). Abdomen was the most common infection site and the admission was mainly surgical, both in S and in NS.

**Table 1:** patients’ characteristics.

| Variable | Survivors | Non-Survivors | p |
| --- | --- | --- | --- |
| N. | 40 | 14 |  |
| AGE (MEDIAN -IQR) | 69 (60.25-73) | 67 (61.25-77.00) | n.s. |
| MALE | 31 | 23 | n.s. |
| SAPS (Mean ± SD) | 41.1 ± 11.7 | 58.3 ± 16.4 | 0.0003 |
| PCT(Median - IQR)<br>r.v. : < 0,5 ng/ml | 48,8 (17,9-93,0) | 8,4 (4,2-22,4) | 0,01 |
| CRP (Median - IQR)<br>r.v. < 5 mg/L | 23,0 (16,0-32,2) | 20,4(14,9-32,9) | n.s. |
| ADMISSION |  |  |  |
| Medical | 5 | 5 | n.s. |
| Surgical | 26 | 5 | n.s. |
| Transferred from other ICU | 1 | 1 | n.s. |
| Emergency Dept | 8 | 3 | n.s. |
| Source of sepsis |  |  |  |
| GI tract | 25 | 5 | n.s. |
| UTI | 8 | 5 | n.s. |
| Skin & soft tissues | 2 | 2 | n.s. |
| Lung | 1 | 1 | n.s. |
| Cancer | 0 | 1 | n.s. |
| Heart | 1 | 0 | n.s. |
| Prosthesis | 1 | 0 | n.s. |
| Unknown | 2 | 0 | n.s. |

Legend: r.v.: reference values; PCT: procalcitonin; CRP: C-reactive protein.

CLI was similar between S and NS both at day 0 and 4 but it significantly decreased at the end of the treatment only in the former group (Table 2).

**Table 2:** Time course of CLI.

| **CLI (median-IQ range)** | **Survivors** | **Non-Survivors** | **p** |
| --- | --- | --- | --- |
| **day 0** | 102 (86-144) | 109 (55-212) | n.s. |
| **day 4** | 53 (21-78) | 64 (22-101) | n.s. |
| **p D0-D4** | <0.001 | 0.051 |  |

The antimicrobial appropriateness did not differ significantly in S and NS.

Ig values before starting eIg infusion were slightly below normal level in both groups, in particular IgM were low in 23% of S vs 22,5% in NS, IgA were low in 7.5% both in S and NS, IgG were low in 46.2% of S and 57.5% of NS.

The time course of the blood values of Ig in survivors and non-survivors is reported in Table 3: at D0, the Ig concentration was similar in survivors and nonsurvivors except for IgA that were significantly higher in NS. At D4, IgG and IgM increased in both groups but only IgA were significantly higher in NS; at D7, IgM were non significantly higher in the S (p = 0.05) whereas IgA were significantly higher in NS (p = 0.02).

**Table 3:** Variation of blood levels of Ig.

| Class | Day | Survivors | Non-Survivors | p |
| --- | --- | --- | --- | --- |
| **IgM**<br>(r.v. 40-230 mg/dl) | 0 | 56 (41-73) | 82 (43-114) | n.s. |
|  | 1 | 94 (74-110) | 105 (58-119) | n.s. |
|  | 4 | 125 (105-150) | 129 (108-138) | n.s. |
|  | 7 | 152 (105-214) | 79 (69-82) | 0.05 |
| **IgA**<br>(r.v. 70-400 mg/dl) | 0 | 153 (116-228) | 245 (180-314) | 0.04 |
|  | 1 | 225 (174-331) | 317 (201-349) | n.s. |
|  | 4 | 252 (214-323) | 438 (304-445) | 0.02 |
|  | 7 | 269 (223-346) | 441 (347-482) | 0.04 |
| **IgG**<br>(r.v. 700-1600 mg/dl) | 0 | 679 (497-874) | 823 (447-955) | n.s. |
|  | 1 | 1221 (950-1376) | 1086 (785-1320) | n.s. |
|  | 4 | 1307 (1122-1699) | 1538 (1248-1774) | n.s. |
|  | 7 | 981 (913-1565) | 1315 (1085-1339) | n.s. |

Legend: r.v.: reference values.

Considering the Ig variation during the days, in D0-D4 and D0-D7 IgM and IgA increased significantly only in S (Figure 1 and 2); the IgG increased but non significantly in both groups (Figure 3).

**Figure 1:**
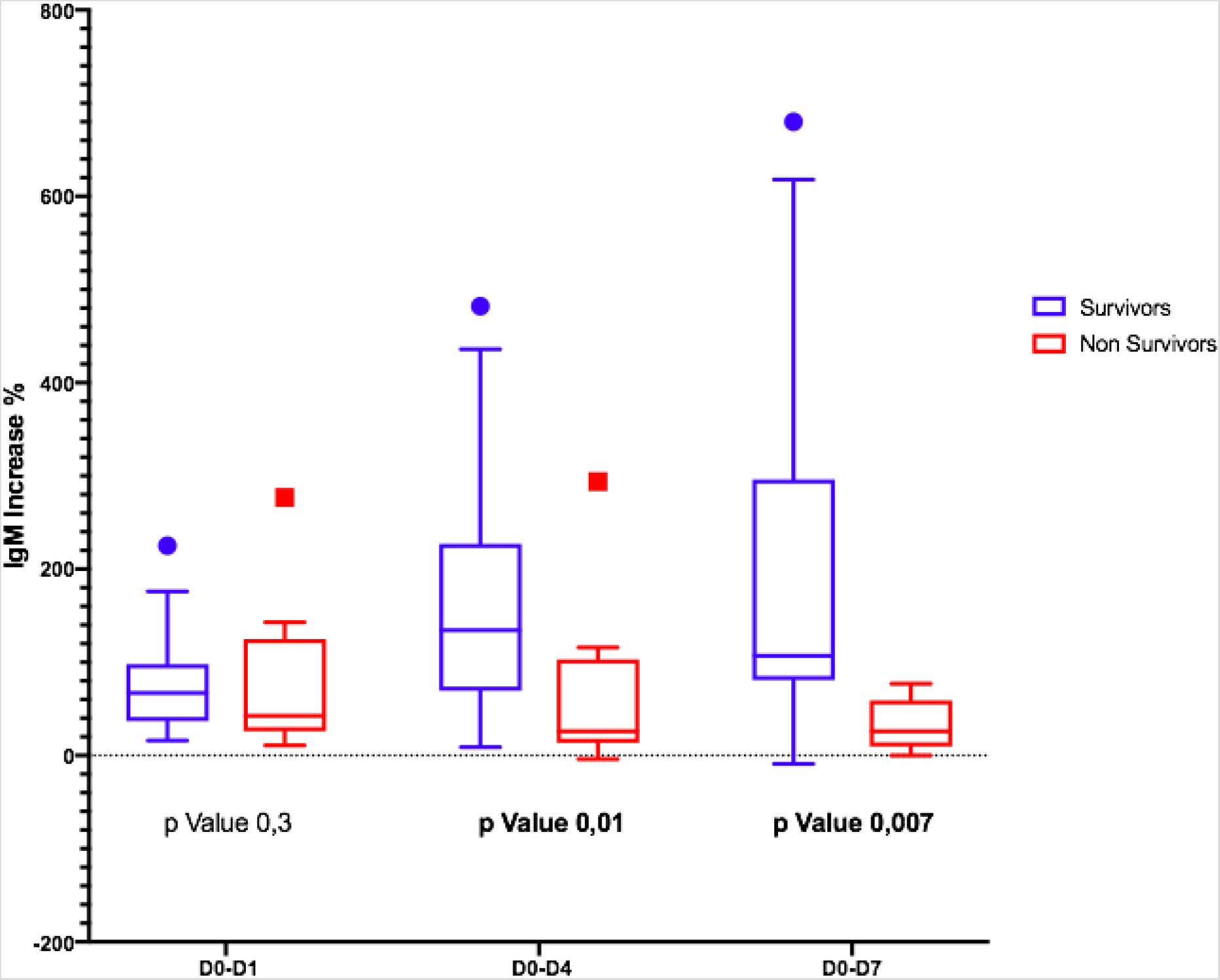
Differences in IgM variation between survivors and non-survivors.

**Figure 2:**
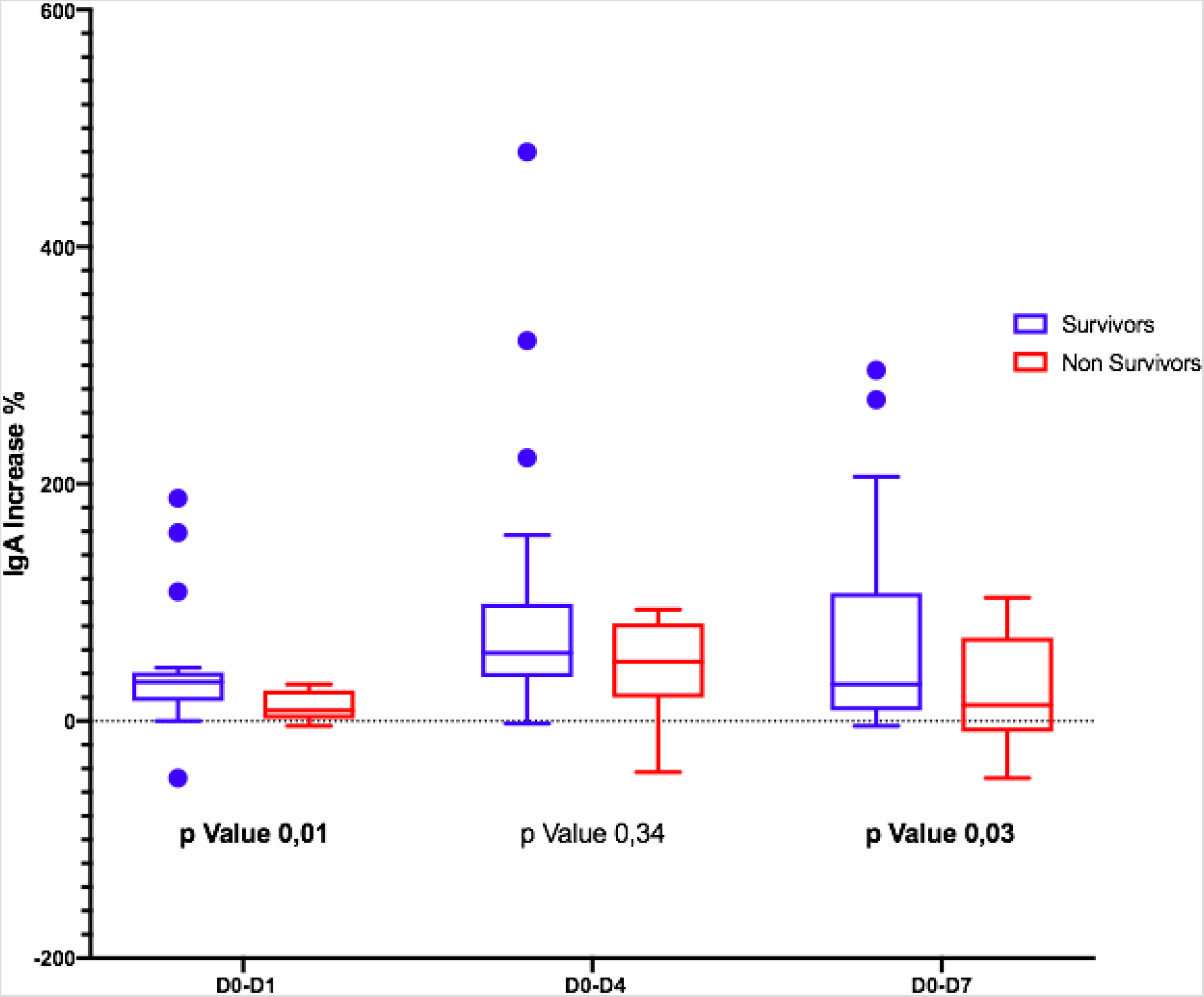
Differences in IgA variation between survivors and non-survivors.

**Figure 3:**
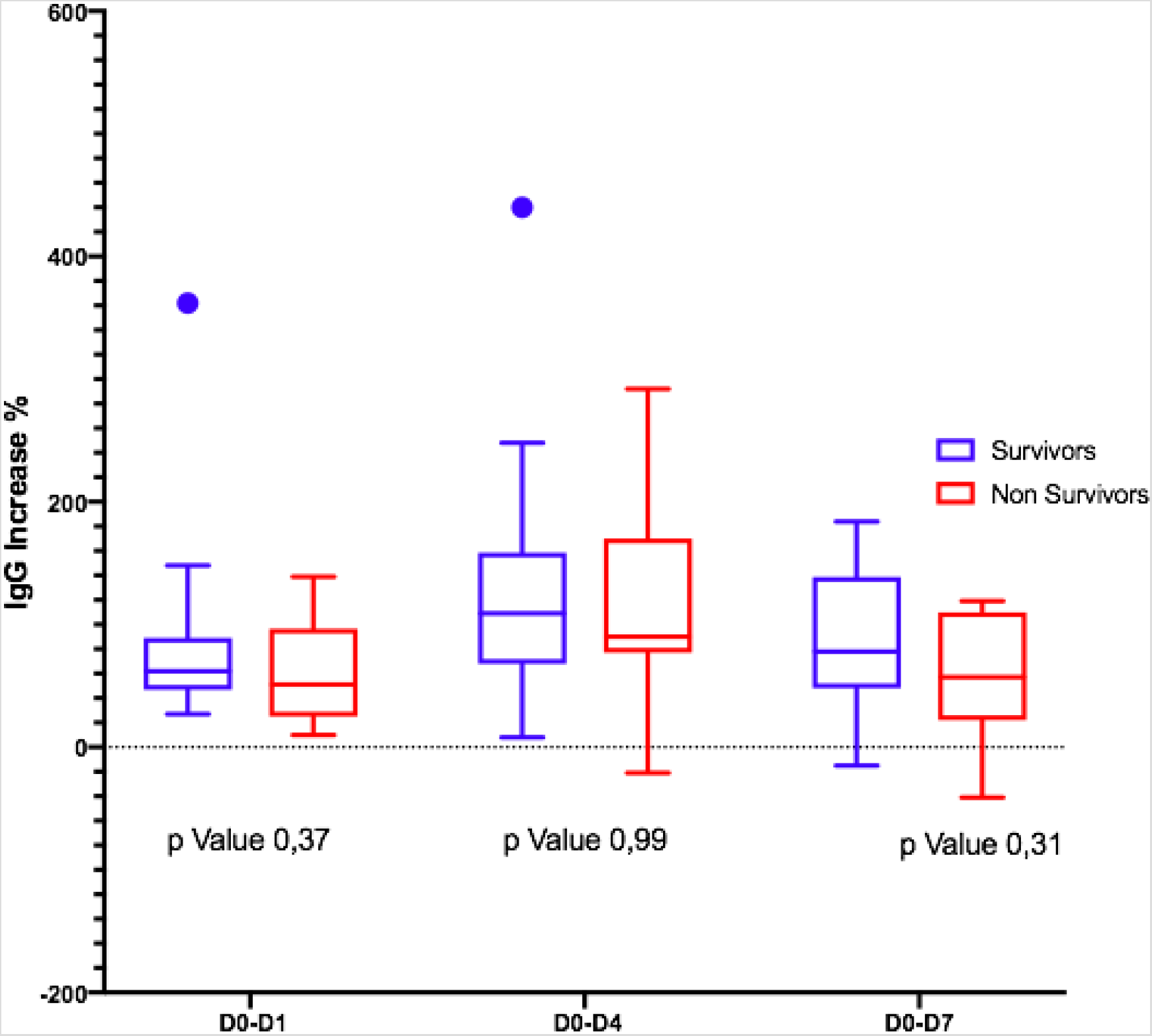
Differences in IgG variation between survivors and non-survivors.

## Discussion

A number of recent observational studies and meta-analysis demonstrated that in septic shock patients the administration of eIg (a) is associated with a higher survival^6–8^, (b) is effective also when MDR strains are involved^6,19^, but (c) is likely time-dependent, as the mortality increases by ∼ 6% for each day of delay following the onset of septic shock even when the antibiotic treatment is appropriate^19,20^. It is conceivable that these results can be ascribed to the enhancement and/or the restoration of patients’ adaptive immune capabilities and, consequently, that a threshold level of native circulating ɤ-globulins could exist below which either the morbidity or mortality of septic patients could increase. Actually, different investigations were addressed toward the relationship between the blood levels of native Ig and the outcome of septic patients, but the results are somewhat controversial: whereas some investigators found that either isolated or combined low levels of IgG, IgM and IgA were associated with a decreased survival^16,21–23)^ and Giomarellos-Bourboulis et al^7^ showed that the progress from severe sepsis to septic shock and death was marked by decreased blood levels of IgM and, other authors reported different results: in a recent meta-analysis Shankar-Hari et al^24^ demonstrated that low levels of IgG and IgM in septic patients were not associated with a poor outcome; the same author found an inverse correlation between low IgG and elevated free light chains λ levels at the ICU admission but these changes did not influence the outcome^25^. Moreover, IgM appear to play a role also in non-septic conditions, as higher levels of this molecule have been found in survivors of a group of non-septic critically ill patients as compared with non-survivors^26^.

Thus, if is the relationship between endogenous Ig and the outcome of patients with septic shock, is far from clear, even less if less is known about the kinetics of ɤ-globulins in patients given IvIg and their possible influence on the survival. Indeed, to clarify the relationship (if any) between the pretreatment levels of γ-globulins, their variations during and after the IvIg administration and the outcome could help to identify the best candidate for this treatment. With this aim, this issue has been studied in two non recent investigations: in the first, performed in septic patients randomized to receive IvIg at the dose of 900 mg/kg of body weight or placebo during 2 days, the blood values of IgG were in the normal range at admission and increased only in the treatment group but without any effect on the mortality; similar results have been reported in the second study which involved cardiac surgery patients developing postoperative sepsis and treated with the same amount of IvIg^27,28^.

Our results obtained in patients given eIg only partly confirm previous investigation but add other pieces of information.

First, NS presented higher initial levels of ɤ-globulins although this difference was significant only for the IgA. In S, the higher PCT at admission likely reflects a more pronounced natural immune response that appears blunted in NS^29,30^.

Second, the trajectory of ɤ-globulins during and immediately thereafter the administration of eIg was different between S and NS: whereas the IgG and the IgA increased in both groups even if less markedly in NS, in the S group at D4 the IgM more than doubled their initial values and almost tripled at D7. This finding contrasts what has been demonstrated in septic shock patients studied along a 4-weeks period by Giamarellos et al^7^ who observed that (a) in S the levels of native IgM sharply increased for the first four days but returned at the baseline values after six days and remained stable till the 28^th^ day; (b) conversely in NS, the IgM presented only minor fluctuations during the four weeks following the onset of sepsis; and, (c) at the 28^th^ days, the IgM levels of both groups overlapped. In our patients, different factors could have been responsible for the higher levels of IgM at D7 in S, including (a) an increased production of endogenous IgM; (b) the reduction of the CLI favoring the retention of these molecules into the bloodstream; and (3) their reduced consumption attributable to the decreased bacterial or PAMP load; (4). Conversely, in NS the ongoing production of endogenous IgG and IgA possibly associated with the reduced production and/or the consumption of IgM in NS.

Different mechanisms can account for the different of time courses of γ-globulins between S and NS, including (a) the ongoing production of endogenous IgG and IgA possibly associated with the reduced production and/or the consumption of IgM in NS; (b) a higher pathogen or PAMP load and the consequent increased opsonization and clearance of the IgM molecule; and (c) the leaking from the bloodstream into the interstitial space of IgM through a more permeable capillary endothelium as suggested by their higher CLI in NS^31,32^.

## Conclusions

We observed a different Ig trajectory between S and NS, which could be related either to (a) different capillary permeability in the two groups and/or (b) higher Ig consumption in NS.

We don’t have enough evidence to set a target Ig value to achieve but, looking at these results, we can try to identify a specific target population that can benefit from higher dosages of eIg, namely those patients whose IgM and IgA values fail to increase after the first three days of treatment.

Due to lack of high quality evidence to support the widespread use of eIg as adjunctive therapy for sepsis, large-scale high-quality RCTs are warranted to confirm the benefits of eIg in the treatment of sepsis syndrome, to define the specific target population and the correct eIg dose.

## Data Availability

all relevant data are available for the rewievers

